# A minimal model for household effects in epidemics

**DOI:** 10.1101/2020.07.09.20150227

**Authors:** Greg Huber, Mason Kamb, Kyle Kawagoe, Lucy M. Li, Boris Veytsman, David Yllanes, Dan Zigmond

## Abstract

Shelter-in-place and other confinement strategies implemented in the current COVID-19 pandemic have created stratified patterns of contacts between people: close contacts within households and more distant contacts between the households. The epidemic transmission dynamics is significantly modified as a consequence.

We introduce a minimal model that incorporates these household effects in the framework of mean-field theory and numerical simulations. We show that the reproduction number *R*_0_ depends on the household size in a surprising way: linearly for relatively small households, and as a square root of size for larger households. We discuss the implications of the findings for the lockdown, test, tracing, and isolation policies.

## 1. Introduction

The rapid spread of the SARS-CoV-2 virus and the COVID-19 pandemic is a threat to global public health with very few precedents. The policy response thus far has often been informed by epidemiological modeling, such as [1]. This paper offers some suggestions for improving such models by accounting for intra-household effects. The role of inside-household transmission for COVID epidemiology has been recognized early [2, 3, 4]. In this work we investigate the combination of relatively fast transmission inside the households and relatively slower transmission between them.

An important feature in epidemiological models is the way the individuals come in contact with each other within the model framework. The simplest assumption is the *panmictic* one: everybody comes in contact with everybody else with equal probability, and thus has a chance to infect everybody else [5, 6, 7]. On the other end of the spectrum are detailed individual-based models where we know exactly who contacts whom at the individual level [8, 9, 10].

The widespread COVID-19 pandemic and resultant partial quarantine measures make the panmictic assumption unrealistic: if confinement works, one hopes to lower the number of contacts outside the immediate surroundings of every individual. On the other hand, detailed models are quite effective at explaining what has happened in the past, but predictions about the future and the results of lockdown policies in the context of these models require detailed assumptions on the mode of future contacts between individuals. These details are knowable in hindsight, but are rather difficult to predict. These considerations suggest a third approach: idealized models that are more realistic than the classical panmictic ones, but less detailed than those based on individual data. In this class of models, we describe the way the population interacts through a limited number of well-defined parameters. These parameters are influenced by policy decisions, so models of this kind can be used to predict the result of specific policies on the spread of contagion. An example of such models is Stroud’s model of influenza spreading in a mixed population [11], where the inhomogeneity was represented by a semi-empirical power law, partially justified by simulations, for the probability of a new infection.

In this paper, we describe the effect of household size on sheltering in place, with application to the current COVID-19 epidemic. We assume that, due to shelter-in-place orders, the contacts between people in different households are uniformly reduced. However, no shelter-in-place policies restrict the interactions between the members of the same household. In fact, these interactions may become much closer and more frequent than before the epidemic. The epidemiological data on COVID-19 in close quarters such as cruise ships [12] suggest very high contagion rates in such situations. Therefore, it makes sense to discuss the spread of infection in a model where people are separated into close-knit groups (“households”) with a high rate of contagion within groups relative to the rate of contagion between groups.

The idea of modeling epidemics in a stratified population is, of course, not new. Approaches similar to ours were employed in [13] to simulate the 1918 flu pandemic and in [14, 15] to study the effect of isolation and vaccination. In [16, 17] the dependence of the spread of infection was obtained in a household model, but with different assumptions from ours, which lead to different predictions.

Usually these approaches rely on many parameters to describe the dynamics of transmission. Here, we instead employ a minimal model that simply introduces within-household transmission in a standard SIR (susceptible-infected-recovered) formalism. This approach allows us to isolate the effect of household sizes from other factors that would affect more detailed descriptions. It is also analytically tractable and presents a phase space that depends on only a few parameters, so it can be thoroughly explored numerically. We are thus able to obtain a simple mathematical law that describes the growth of the epidemic’s rate of spread as a function of the household size. While more finely grained models could offer a more detailed description, we expect this law to be universal as long as our main premises —high transmission rate for close contacts and high rate of asymptomatic transmission— are satisfied.

Below, in Section 2, we describe the model in detail and calculate the fast-growing modes. This analysis provides insight into the initial dynamics of epidemics and the influence of various parameters on the rate of infection. In Section 3, we report on the results of simulations and numerical solution of the mean-field equations. In Section 4, we discuss the results and their meaning for lockdown strategies. The conclusions are stated in Section 5.

## 2. Mean-field models

Let us discuss a population separated into households with high contact rates within a household and low contact rates between different households.

We will use the SIR model with three states for each individual: Susceptible, Infected, and Recovered, as shown on Figure 2. The state of a household is described by the number of individuals in each state (*s, i, r*). If *H* is the household size, then *s* + *i* + *r* = *H*. In this section, we use a simple model where the first infection quickly leads to the whole household being infected, so the simplified state diagram becomes the one shown in Figure 3. Let *S* be the number of households with noone infected, *F* be the number of households with one person infected, and *G* be the number of households with everyone infected. The total number of infected persons is *F* + *HG*. Let us neglect secondary outside transmission, *i*.*e*., the situation when a member of an already infected household gets infected through an outside contact. Then the probability that an arbitrary contact of an infected person is susceptible is *SH/N*, where *N* is the total number of persons. If *β* is the transmission rate, then the number of households is governed by the equations

**Figure 1.**
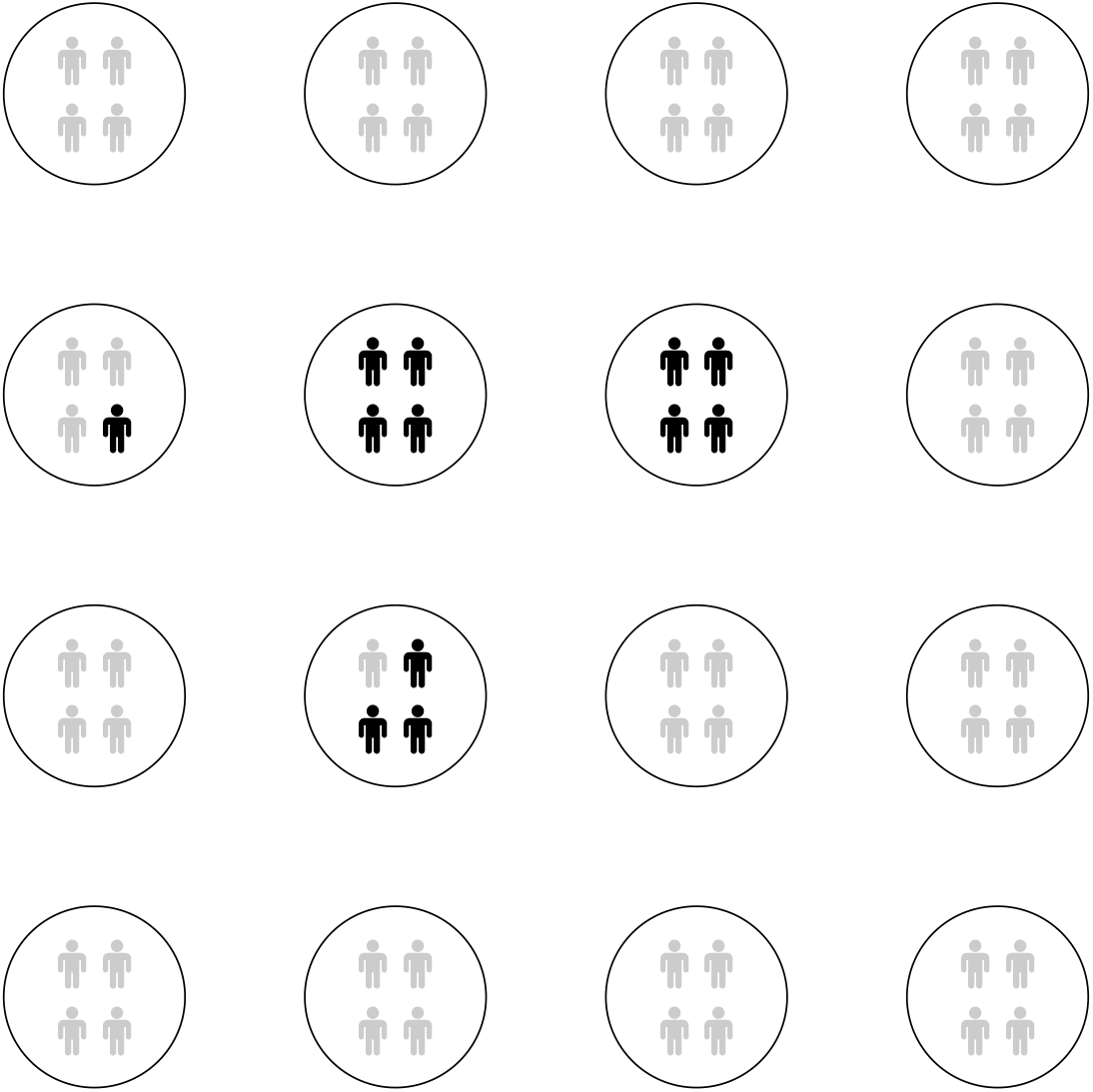
Household model. Black symbols correspond to infected individuals.

**Figure 2.**
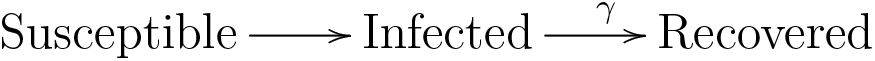
State diagram for an individual in the simplest model.

**Figure 3.**
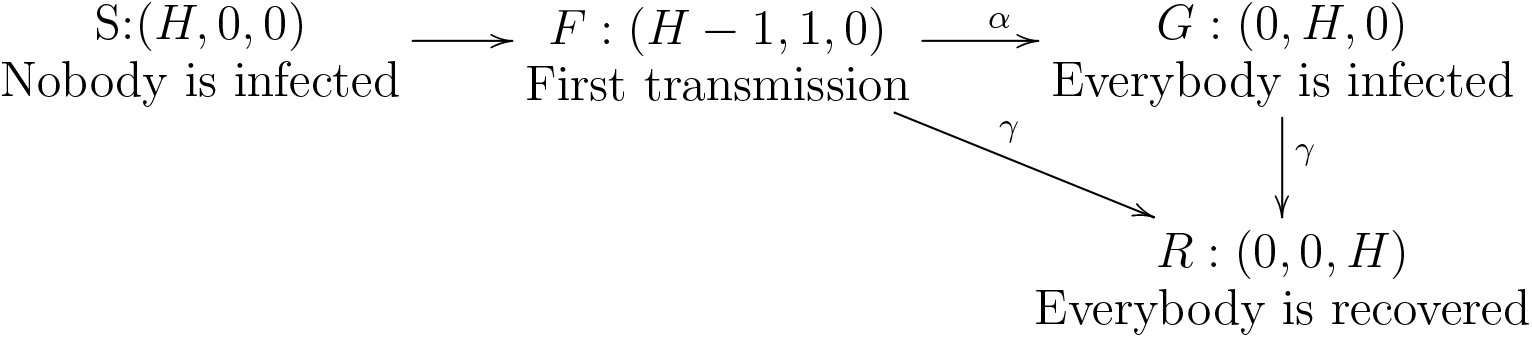
Simplified state diagram for a household. The household state is (*s, i, r*): susceptible, infected, recovered.

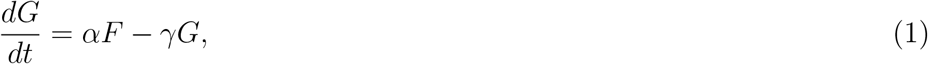

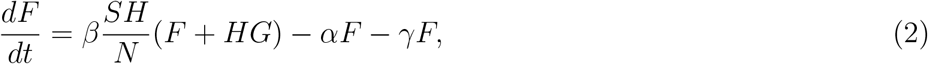

where *α* is the rate of having the whole household infected, and *γ* is the recovery rate. We assume the recovery rate for all household members is the same and neglect the difference in the time of infections of different household members.

If all members of the household are quickly infected by the index patient, the rate constant *α* does not depend on the household size *H*. If contagion spreads through aerosols or shared surfaces [18], the rate of spread does not depend on *H*. This suggests that the dependence *α*(*H*) is weak or non-existent. Therefore, in the subsequent models we neglect this dependence, and discuss its possible effects in the Appendix.

In the early stages of the epidemic, far from saturation, *SH ≈ N*, and equations (1) and (2) are linear. The positive eigenvalue of the system corresponds to the growing mode and is equal to

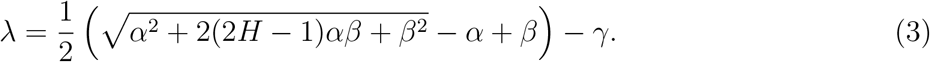

A fast in-household transmission corresponds to *α* ≫ *β*. Equation (3) shows that there are two regimes for the transmission. For small household sizes, when 2(2*H* − 1)*β* ≪ *α*, the exponent for the growth of infections depends on *H* linearly, as *λ ≈ Hβ* − *γ*. However, for larger households, when 2(2*H* − 1)*β* ≫ *α*, the effective growth rate is proportional to the square root of *H*, 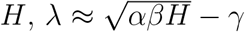. This finding is in contrast with the one in [16], where a linear dependence on *H* was found under different assumptions.

It is customary to quantify epidemic-growth potential using the basic reproduction number *R*_0_, which is equal to the increase in the number of infections during the course of disease for an individual patient. For this model

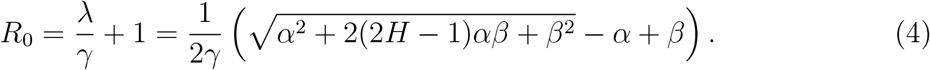

We will assume *γ ≈* 0.125 days^−1^ and the rate of spread in the absence of lockdown *β ≈* 0.3 [19]. Setting different values for *β* under lockdown, we plot the values of *R*_0_ on Figure 4.

**Figure 4.**
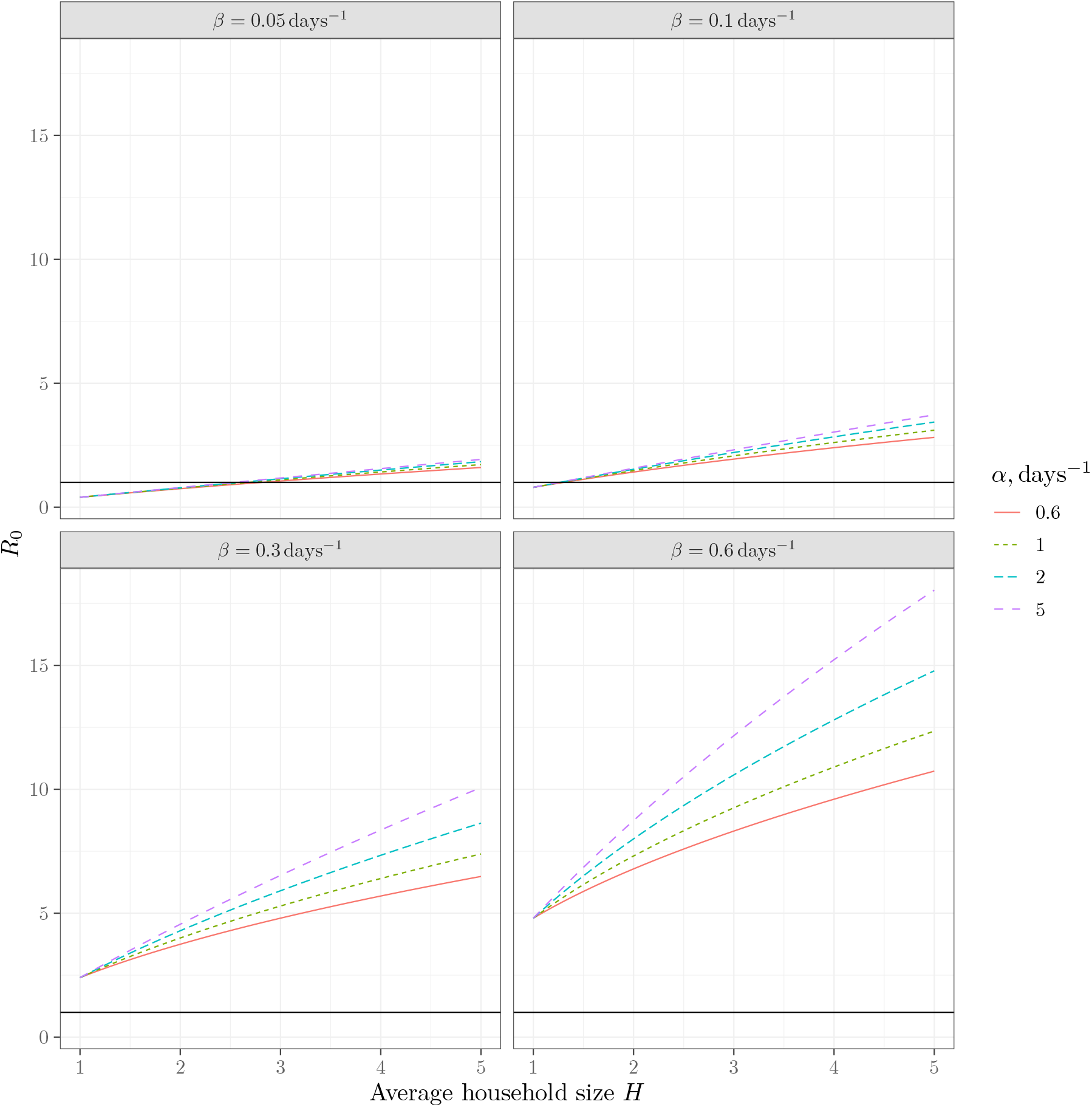
*R*_0_ for the model with *γ* = 0.125 day^−1^. The horizontal line corresponds to *R*_0_ = 1. Each subfigure corresponds to a fixed *β* with a curve for each value of *α*.

## 3. Numerical modeling

Eigenvalue analysis of equations (1) and (2) is applicable in the linear regime only. For the situation where the depletion of the susceptible population is non-negligible, we need either to solve these equations numerically or perform simulations. Below we report the results of both approaches.

We performed numerical simulations to validate the analytical models of household infections described in the above section. In each scenario that we studied, we took a population of *N* = 200 000 individuals, placed each individual into a house for a range of different household sizes, and subsequently initialized a small random subset of 20 individuals in the population with an infection. For each step, we divided the infection into two phases:

i. Panmictic phase: each infected individual can infect any uninfected individual in the simulation, with a probability 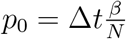, where *β* is the daily rate of infection in the panmictic phase, *N* is the population size and Δ*t* is the time step of the simulation.
ii. Household phase: each infected individual can infect any uninfected individual in their house, with a probability Δ*tα*, where *α* is the daily rate of infection inside the household.

We studied these regimes using Δ*t* = 0.1 and *γ* = 0.125. We ran a different simulation for various combinations of *α, β* and *H*. For each combination of values we ran the simulations *n* = 10 times. For the same parameters we numerically solved equations (1) and (2). The results are shown in Figure 5.

**Figure 5.**
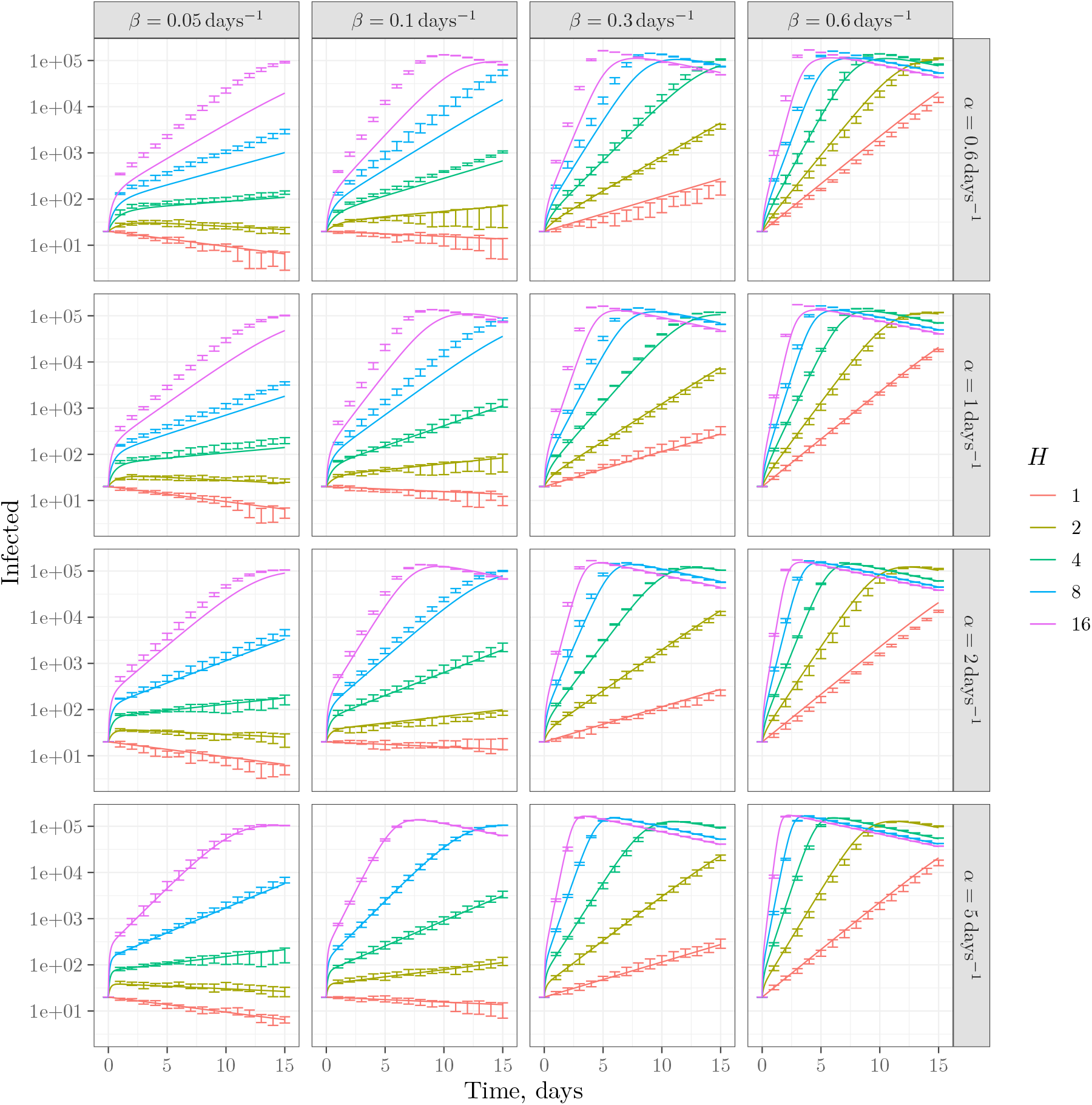
Simulations and mean-field predictions. The lines correspond to numerical solution of equations (1) and (2). The error bars correspond to the confidence intervals for repeated simulations computed as 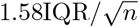 [20]. Each subfigure corresponds to a combination of *α* and *β* as shown on the panels with a curve for each value of *H*.

The model shows a very good agreement between the predictions of the mean-field theory and numerical experiments, especially for large in-household transmission *α*. It should be noted that the meaning of *α* is subtly different for simulations and mean-field theory. For simulations, *α* is the rate for an individual to become infected inside a household; while for mean-field theory, *α* is the rate for a household as a whole to become fully infected. These two definitions are expected to be convergent in the limit of fast inside infections, as seems to be the case from Figure 5.

It is interesting to look at the total number of cases during the epidemics. The results of our simulations for *N* = 200 000 are shown on Figure 6.

**Figure 6.**
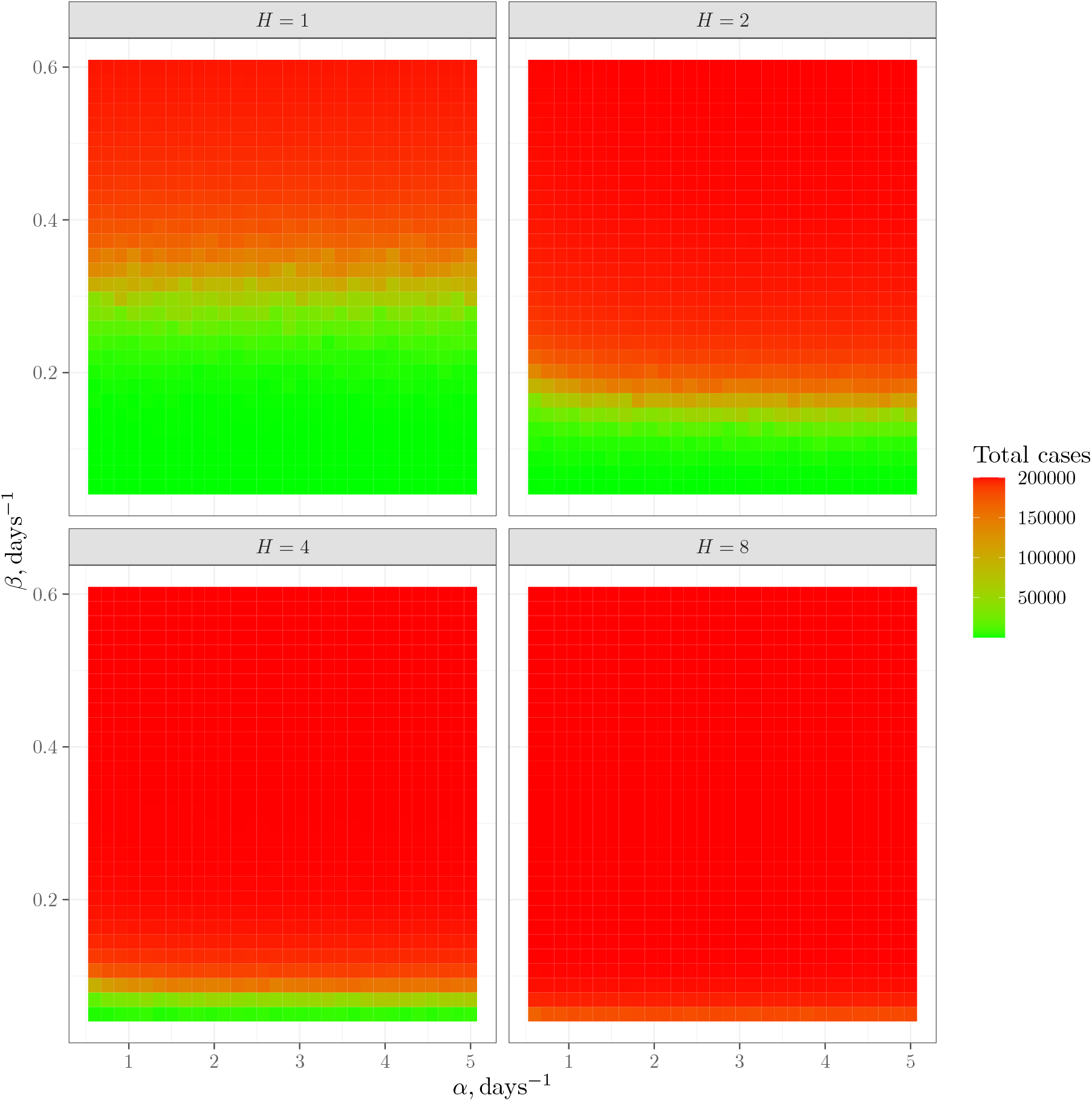
The total number of cases for simulations with *N* = 200 000. Each subfigure corresponds to a value of *H*.

## 4 Discussion

Our approach is based on the existence of two time scales: fast transmission inside households and slow transmission between the households. Following the scaling approach (see, for example, [21]), we can imagine the households being “super-individuals” with renormalized interaction parameters. Since, in this model, *H* people have *H* times as many contacts as one person, one can naïvely expect that the spread of contagion scales as the household size *H*. However, as confirmed by mean-field calculations and numerical simulations, the scaling law is less steep: the spread grows only as 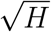. This fact may have profound policy implications, suggesting the possible establishment of larger “households” such as social circles) rather than a uniform reintegration of all contacts during reopening. However, further modeling and comparison with empirical data would be necessary before implementing the results of this paper as policy. A rather strong dependence of the total number of cases over the time span of epidemic on the household size (Figure 6) stresses the necessity of caution at reopening.

The assumptions underlying the model reveal its natural limitations. For mean-field calculations (but *not* numerical simulations), we assumed that inside-household dynamics are much faster than the transmission between households. This allowed us to neglect the details of transmission inside households and use an idealized model depending just on the mean time between the first infection and the completely infected household. This also allowed us to neglect secondary between-household infections (*i*.*e*., household members of an infected individual getting infected by their outside contacts rather than by the inside ones). Our numerical simulations did not employ this assumption explicitly. However, by selecting a very simple inside-household infection model (pair interactions between household members) we made the assumption implicitly: the details of the inside transmission should not matter if the transmission is fast. Another assumption for the mean-field model is the neglect of random fluctuations: we used mean values to construct the equations. We also neglected the dependence of the time for a household to become infected on the household size. This dependence should be significant for very large “households” (see Appendix).

While we use the word “households” throughout this paper, our model may describe situations beyond the simple picture of families waiting out the epidemics together. A dormitory, cruise ship, or even apartment building with a common ventilation system may represent examples of a large “household” (see [22] on the role of common ventilation systems). While in the calculations here we assume “households” of equal size, the model could be generalized for a mixture of different groups with a high level of transmission inside groups and a low level of transmission between the groups.

Another possible application of our model is related to the observation that one of the ways to slowly reopen may involve the relaxing of social distancing within certain groups while maintaining it between the groups. In this case, these “relaxed” groups can be treated as “super-households”. An important application of the super-household concept could be a rudimentary testing-and-tracing policy, in which we test all members of the super-household of any asymptomatic infected individual, instead of a more expensive attempt to trace and test all their contacts. This application is important because asymptomatic transmission seems to be one of the salient and dangerous features of the current COVID-19 epidemic [23].

In subsequent papers, we will analyze testing-and-tracing strategies for the household-based shelter-in-place policy.

## 5 Conclusions

We studied epidemic spread in a stratified population, such as under shelter-in-place orders, using mean-field theories and numerical simulations. We found that:

i. Under shelter-in-place conditions the spread of the epidemics depends on the size of the “household” (*i*.*e*., the group of people sheltered together).
ii. The dependence of the rate of spread on the household size is linear for relatively small households and a square-root law for larger households.

We used idealized models to study the general features of the epidemics in the shelter-in-place conditions. More realistic models may add to these findings, but we expect the main results above to hold.

## Data Availability

Simulation codes are available upon request.

## Acknowledgements

GH, LML, and DY are supported by the Chan Zuckerberg Biohub. The authors are grateful to Daniel S. Fisher, who suggested an elegant formalism for the analytic model, and to Rob Phillips for his inspiration and for including us in his stimulating COVID discussion group.

## Appendix A. Dependence of *α* on *H* for large households

In models with small households, *α* has very little dependence on *H* since most household members will be infected in short order by the household’s first infected individual. In this case, we ignore secondary infections. However, in large households, it may be required to take secondary infections within a household into account.

Here, we look at the dependence of *α* on *H* in a regime where secondary interactions are relevant. In particular, this model uses the same method of in-house infection as the numerical simulations.

Assume that the infection rate within a household is proportional to the number of “connections” between infecteds and susceptibles. In a household of size *H >* 1 with *n* infecteds, we may draw *H*(*H* − 1)*/*2 connections between household members; *n*(*H* −*n*) of those connections will be between infecteds and susceptibles. That is to say, the infection rate is *α*_*n*_ = *α*_0_*n*(*H* − *n*) for some constant *α*_0_ which is independent of *H* and *n*.

The total time to infect the entire household starting with one infected is therefore

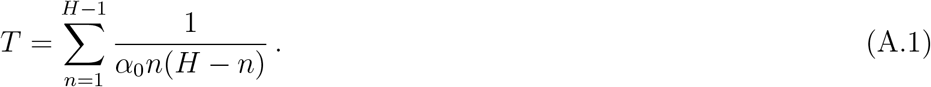

We now reduce this expression for *T*.

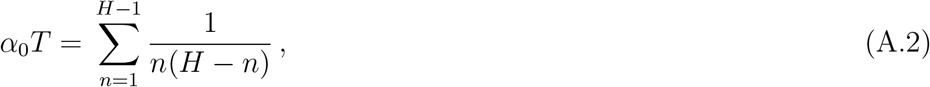

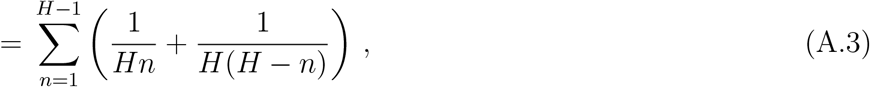

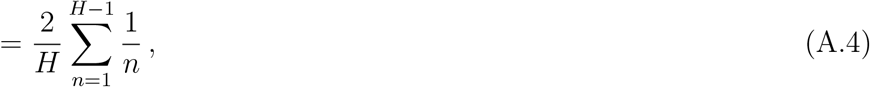

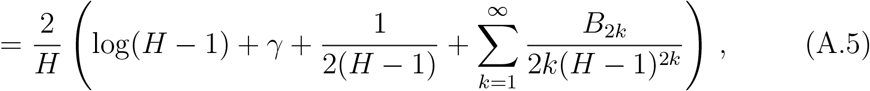

where *γ ≈* 0.577 is the Euler-Mascheroni constant and *B*_*m*_ is the *m*th Bernoulli number. Although it is not obvious that the terms after *γ* become vanishingly small for large *H* (since the Bernoulli numbers grow quickly), it is well known that these terms are only a small correction in this expression for the harmonic series. Therefore, defining *α* = 1*/T*, we may write the approximate expression

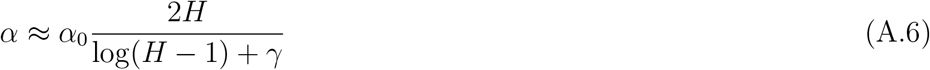

for *H >* 1.

## Notes

### Competing Interest Statement

The authors have declared no competing interest.

### Author Declarations

Theoretical modelling only, no such oversight required.

